# Combining Indoor Positioning Using Wi-Fi Round-trip Time with Dust Measurement in the Field of Occupational Health

**DOI:** 10.1101/2021.04.14.21254456

**Authors:** Hajime Ando, Shingo Sekoguchi, Kazunori Ikegami, Hidetaka Yoshitake, Hiroka Baba, Toshihiko Myojo, Akira Ogami

## Abstract

**[Objectives]:** The measurement of personal exposure has been attracting increased attention for the control of hazardous substances. Although the combination of location information and personal exposure is considered highly useful, to-this-date, no simple or highly accurate indoor positioning method has been presented. Recently, Wi-Fi round-trip time (RTT) has attracted scientific attention as a highly accurate indoor positioning method. In this study, we investigated the possibility of the use of a combination of this indoor positioning method and small-dust sensors.

**[Methods]:** We measured indoor positioning and dust concentration three times during a walk in a simulated factory. A wearable particle monitor was worn on both upper arms, and a smartphone was attached to the helmet for positioning. An ultrasonic humidifier was used to spray sodium chloride solution in the factory. The measurements were conducted three times on different routes on foot (Experiment A, B, C).

**[Results]:** In indoor positioning, the error percentages, i.e., measurements that were outside the correct measurement area were 7% (49 s/700 s) in Experiment A, 2.3% (15 s/660 s) in Experiment B, and 7.8% (50 s/645 s) in Experiment C. The dust measurements were also taken without interruption. A heat map was created based on the results of both measurements.

**[Conclusions]:** Wi-Fi RTT was able to implement indoor positioning with high accuracy. Thus, the combination of indoor positioning and various sensors can be considered useful for occupational health.

## Background

In Japan, dust exposure is assessed by working environment measurements. There are some problems with this method, such as the fact that it does not always reflect the exposure status of workers when they work while moving. Therefore, an amendment to the Working Environment Measurement Standards has been planned to allow personal exposure measurements instead of conventional working environment measurements for some hazardous substances. Personal exposure monitoring is, thus, expected to be used at workplaces where it has been difficult to measure hazardous substances, such as locations where a) temporary or short-term work was conducted, b) source of dust was moving, or c) where work occurred outdoors. Although dust is not subject to personal exposure measurement by law, recent advances in sensor technology have led to the release of compact PM2.5 sensors. These sensors were originally developed for the measurement of air quality, but we believe that they can also be used for the evaluation of personal exposure to dust. Therefore, we developed a wearable particle monitor (WPM), which uses a commercial sensor (HPMA115C0-003, Honeywell International, Inc., New Jersey). Even though the sensor was not calibrated based on strict protocols, the sensor’s module (manufactured by Honeywell) has been reported to have a certain level of accuracy.^1^ Given that this product has not been officially calibrated, it cannot be used for working environment measurements.

When conducting personal exposure measurements, it is necessary to record actions and observations.^2^ However, recording accurate records and observations while conducting measurements is not easy, and requires the assignment of a full-time observer. Given that the details of the work may correspond to the location of the worker, recording the location information of the worker may help the recording of the work. Global Navigation Satellite System (GNSS), represented by the Global Positioning System (GPS), is the standard technology for outdoor positioning. However, for indoor positioning, there is no standard technology such as the GNSS for outdoor positioning because the satellite signal cannot be received indoors. Instead, there are several nonstandard methods, such as Bluetooth low-energy beacons,^3, 4, 5^ Wi-Fi signals,^4, 6^ and various sensor devices.^4^ However, these methods have problems in terms of accuracy, and various measures have been proposed to improve their accuracy characteristics. In 2016, the Wi-Fi round-trip time (RTT) distance measurement method was standardized to IEEE802.11mc as a method with higher accuracy than these methods.^7^ Using this standard method, distance can be measured with a high accuracy by calculating the distance from the arrival time of radio waves, instead of estimating the distance from the radio wave strength, as in the conventional methods. The Wi-Fi RTT protocol enables positioning without synchronizing the clocks of the two parties, thus eliminating the need to use high-precision clocks, such as cesium atomic clocks, and it can be easily installed by using consumer Wi-Fi access points (APs). There are only a few reports on positioning using this standard, but these have been mostly restricted in the field of engineering. Therefore, in this study, we investigated the utilization of Wi-Fi RTT and WPM in the field of occupational health based on the use of a combination of Wi-Fi RTT and WPM at industrial sites.

## Methods

This experiment was conducted in August 2020 at a multipurpose simulated factory at the University of Occupational and Environmental Health, Japan. The overall layout is illustrated in Figure 1.

**Figure 1.**
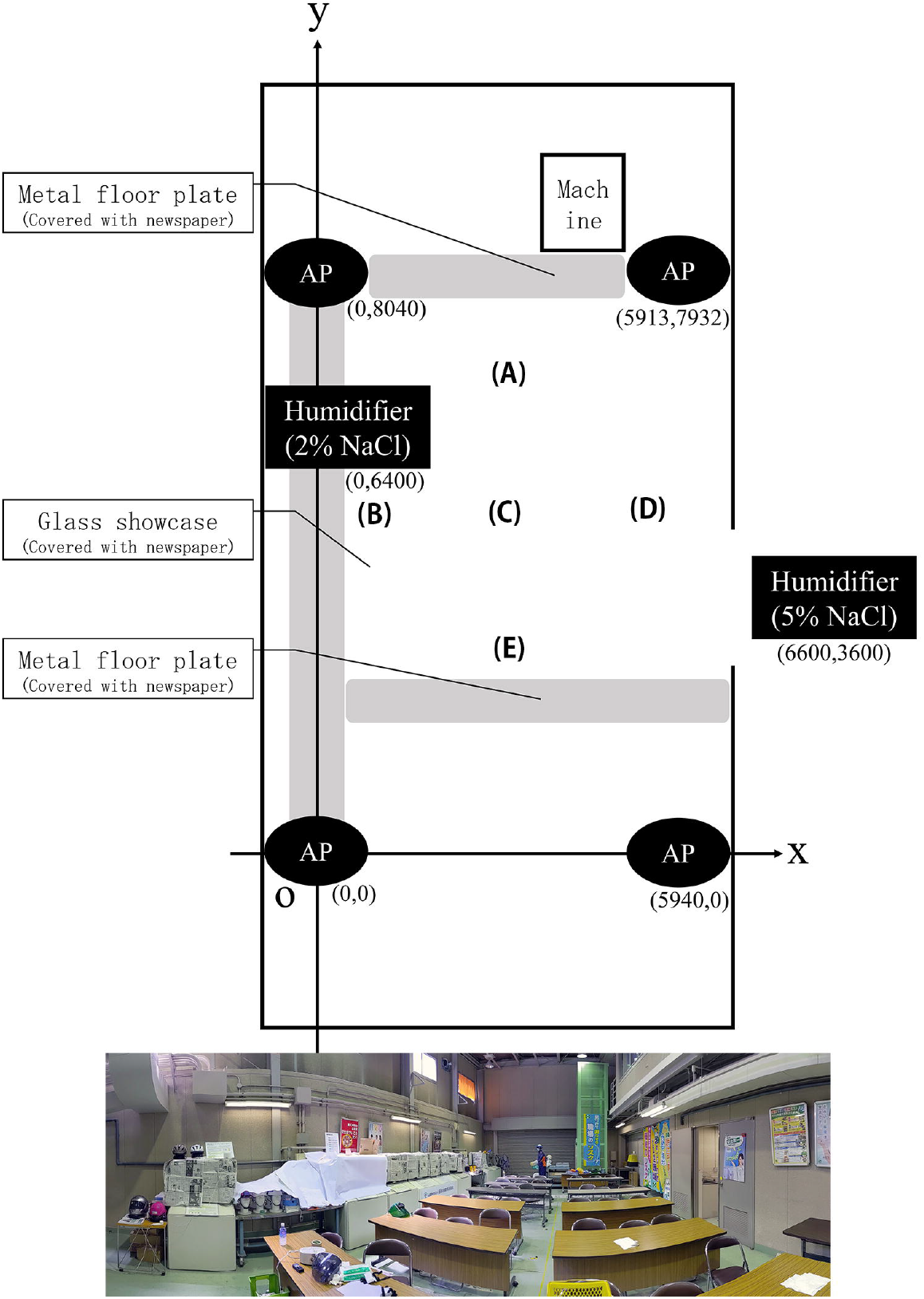
Overview and photographs of the model factory. A to E indicate the positions of the measurement points for accuracy verification. The coordinates measured by the laser rangefinder are A (2900, 6500), B (1900, 3000), C (3000, 3200), D (5700, 2800), and E (3100, 800) (AP, access point).

### Indoor positioning

Indoor positioning was measured using Wi-Fi RTT. We used a Pixel 4 smartphone (Google LLC), a Nest Wi-Fi (Google LLC) router, and three expansion points as APs. Regarding measurement applications, we were unable to find any existing software that matched the requirements of this study. Therefore, we used an open source (MIT License)^8^ software developed by Mr. Darryn^9^ with our own modifications. The major modifications included the change of the coordinate system of the AP, and the addition of a function to save the log. Moreover, we used the developer mode to overcome the limitation of the number of Wi-Fi scans by the Android OS. The coordinates of the AP locations were determined based on measurements with a GLM 50 C Professional (Robert Bosch GmbH, Gerlingen, Germany). The heights of the AP locations were approximately 1500 mm from the floor. Given that the height was the same for all the APs, the Z coordinate was ignored in this study. The APs were placed as shown in Figure 1. The preliminary experiments determined the positioning to be inaccurate in the area around the glass showcase and the area where the floor material was made of steel. Hence, these areas were covered with paper or cloth. In a previous study, it was reported that the accuracy of the positioning varied depending on the direction of movement because of the body which acted as a shield.^10^ Therefore, a selfie stick was fixed to the helmet, and a smartphone was installed. A photograph of the measurer is shown in Figure 2.

**Figure 2.**
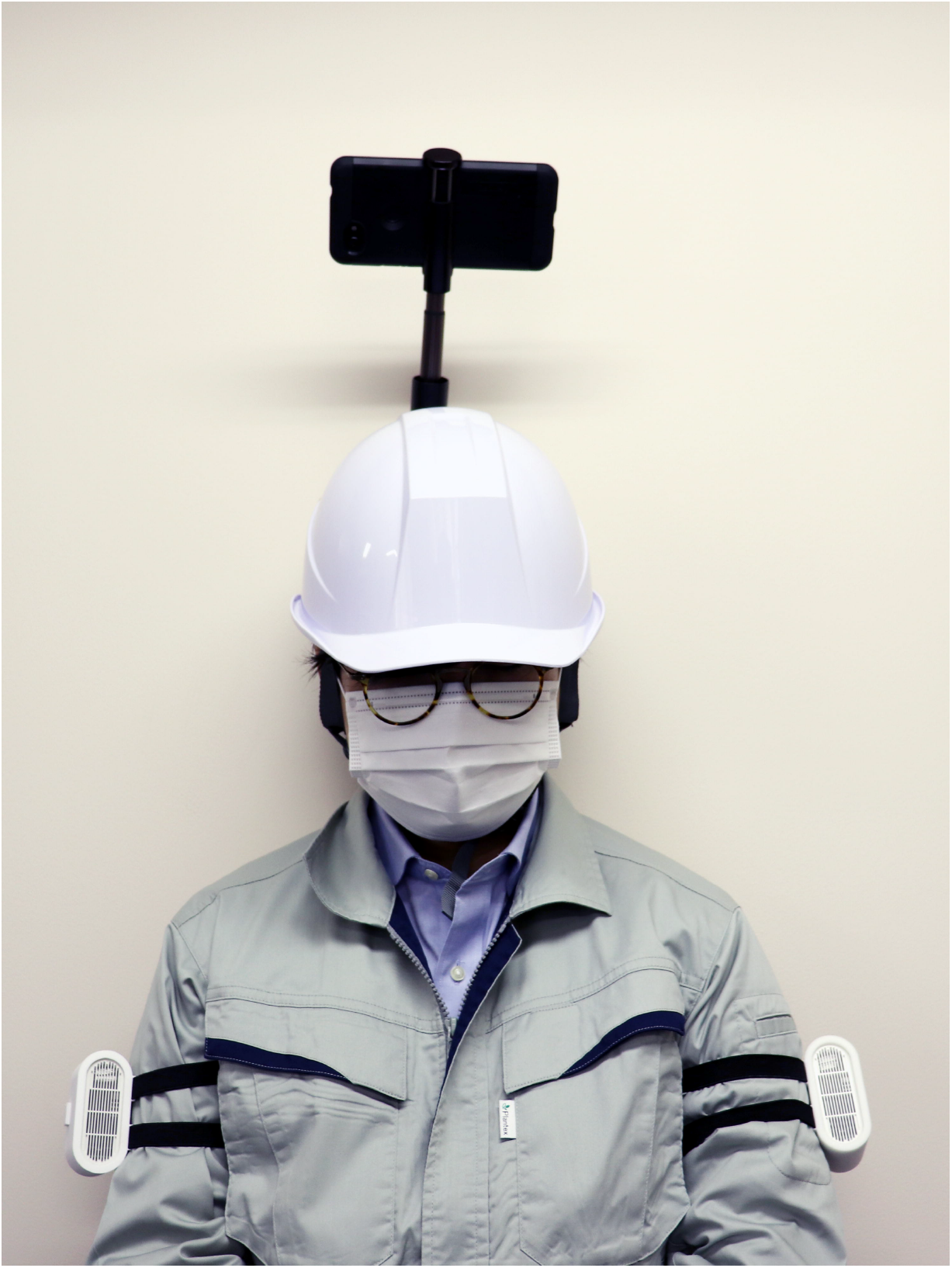
Photograph of the measurer. A wearable particle monitor was worn on both upper arms. A smartphone for indoor positioning was attached to the helmet using a selfie stick.

### Confirmation of measurement accuracy

Prior to this experiment, we checked the measurement accuracy with the use of a with a smartphone which was placed on a helmet and a selfie stick for Wi-Fi RTT measurements at all point locations from points A to E in Figure 1. Measurements were acquired at each point for one minute each. The coordinates measured by the laser rangefinder were used as the standard, and the deviation of the measurement by Wi-Fi RTT was calculated.

### Dust measurement

The WPM was used for the dust measurements. One unit of the WPM was fixed to the left and right upper arms of the person who performed the measurement. Given that there was no dust source in the simulated factory, two ultrasonic humidifiers were installed, and a sodium chloride solution (2% and 5%) was sprayed. Commercial salt was used as the solute, and pure water was used as the solvent. Measurements were acquired every second, and cases with zero readings or no records were excluded. After ventilating the room for each measurement, the humidifier was turned on, and measurements started after the elapse of (at least) 3 min.

### Measurement procedure

Measurements were acquired three times with the use of different routes. The measurer walked the designated route at a low speed of approximately 7.62 m/min. Figure 3 shows the walking route at the time of measurement.

**Figure 3.**
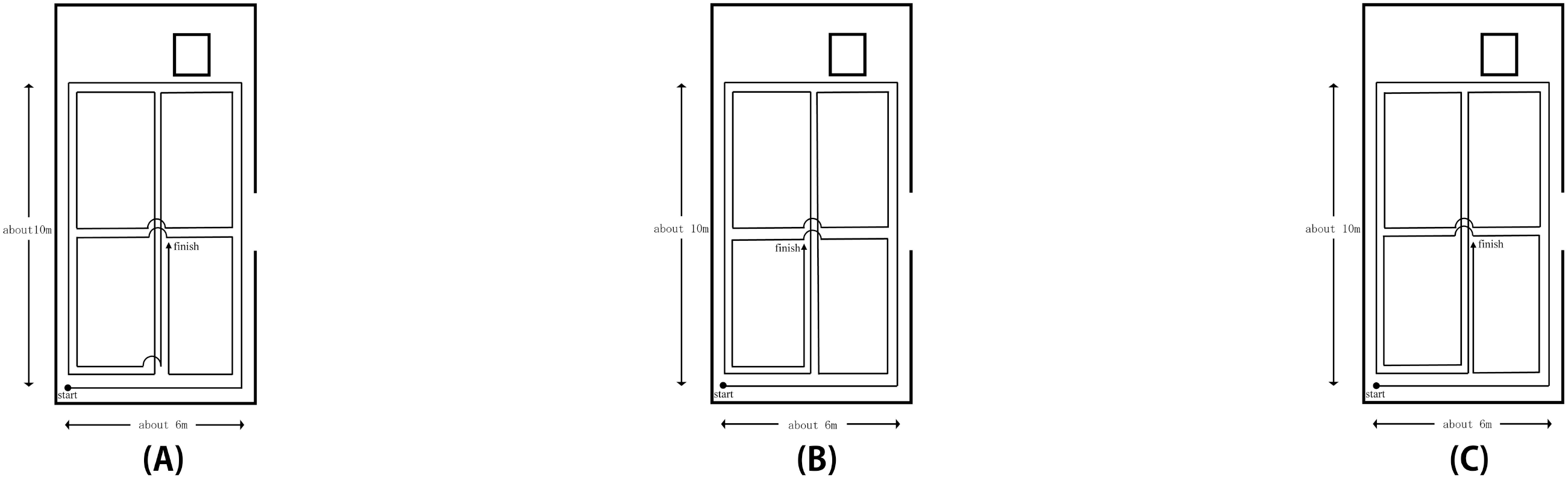
Walking path during the measurement.

### Creation of heat maps

A heat map was created from the positioning information using Wi-Fi RTT and the results of dust measurements by the WPM. The location information was calculated as the median value of the X-and Y-axis coordinates every second. The heat map was created by excluding the coordinate values outside the original circumference range (a rectangular range consisted of four points: (0, −1000), (6000, −1000), (0, 9000), and (6000, 9000)). The dust concentration was determined by using the median of the measured values in the area. The heat map was created using Origin 2020 (64-bit, SR1 Ver9.7.0.188, OriginLab Corporation, Northampton, Massachusetts, USA).

## Results

The results of the accuracy verification are shown in Figure 4. The mean deviation (SD) from the laser rangefinder position at each point was 1087.9 (133.7) mm at point A, 1000.6 (63.7) mm at point B, 748.3 (31.5) mm at point C, 669.2 (94.3) mm at point D, and 918.4 (396.8) mm at point E. The results of indoor positioning are shown in Figure 5. In the locations where the measurer could not exist, such as behind the showcase or behind the wall, which were “outside of the rectangle which comprised the four points of (0, −1000), (6000, −1000), (0, 9000), (6000, 9000),” the error percentages were recorded at 7% (49 s /700 s) in Experiment A, 2.3% (15 s /660 s) in Experiment B, and 7.8% (50 s/645 s) in Experiment C. The results of the WPM measurements are shown in Appendix 1. Figure 6 shows the heat map created by combining the two datasets.

**Figure 4.**
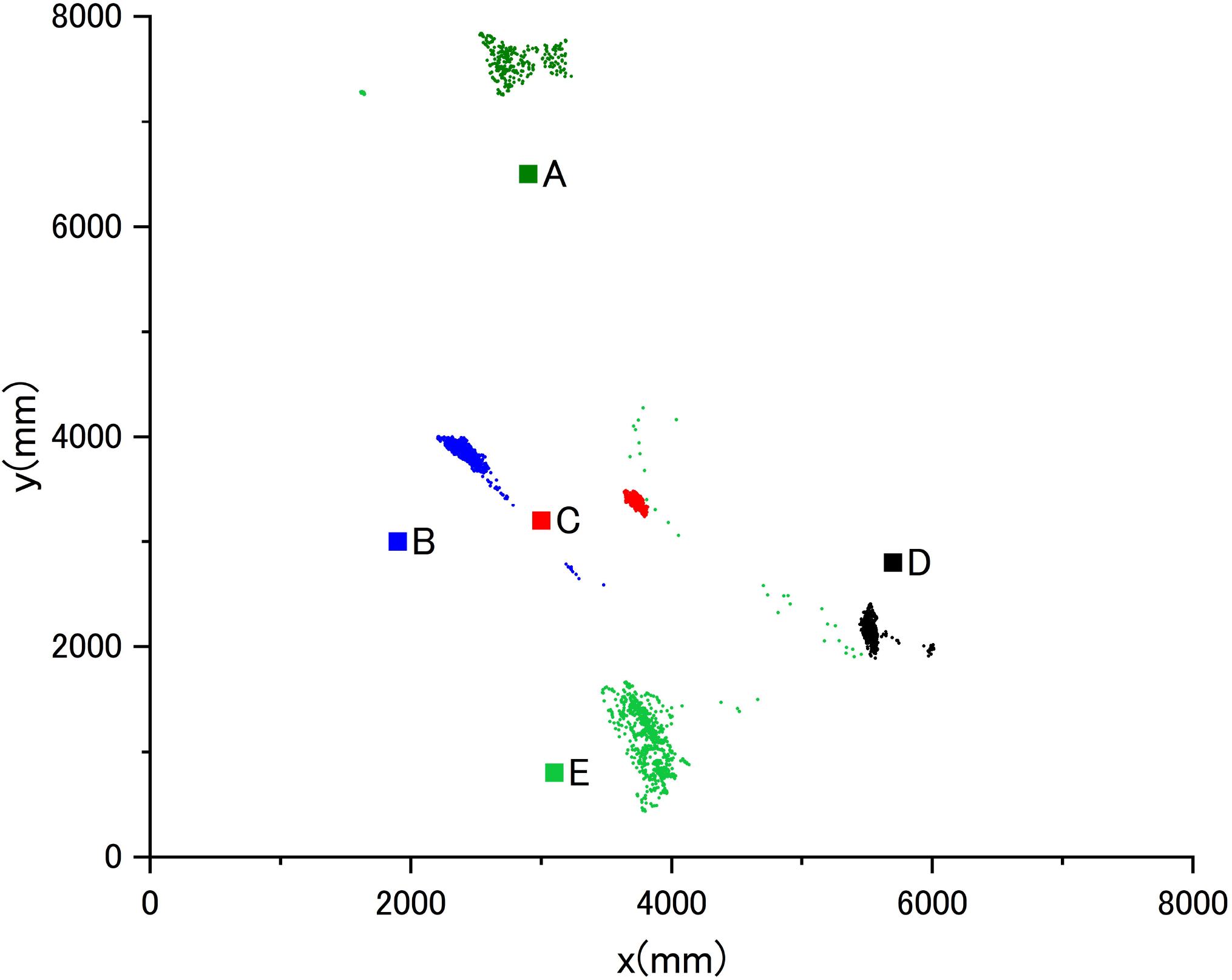
Results of accuracy verification The small dots indicate the position measured by the Wi-Fi RTT at each measurement point. The square dots indicate the actual position of each measurement point.

**Figure 5.**
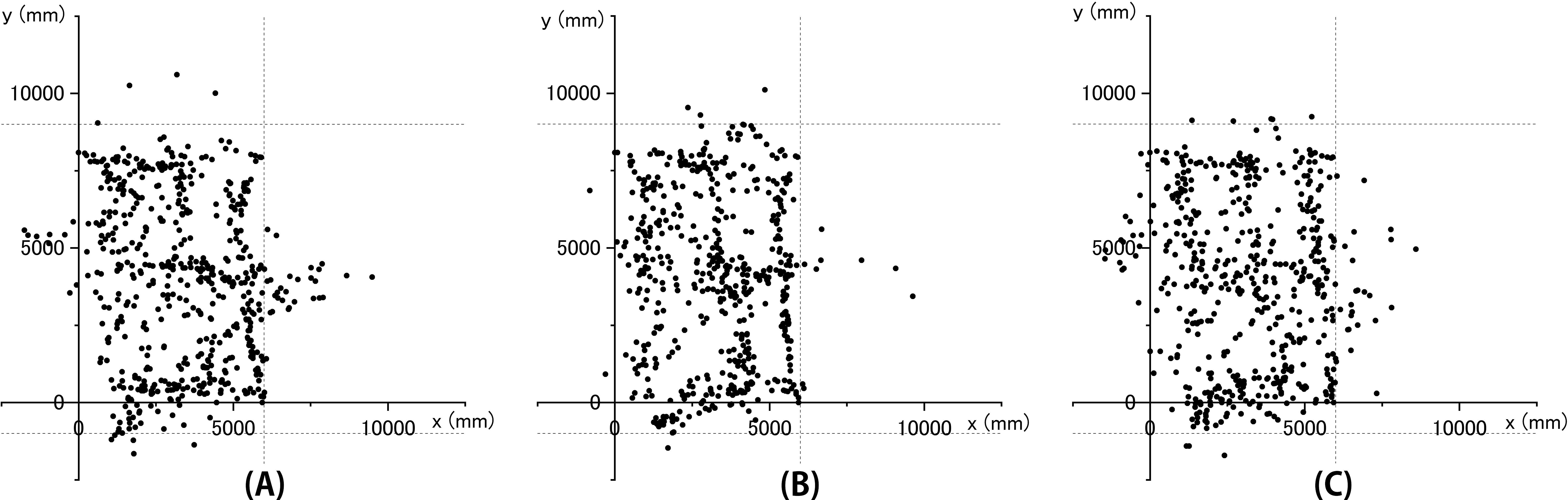
Indoor positioning results. Each dot indicates the position coordinates measured by the Wi-FI RTT. The location information was calculated as the median value of the X- and Y-axis coordinates every second. The range enclosed by the dotted line and the Y-axis is the walking range.

**Figure 6.**
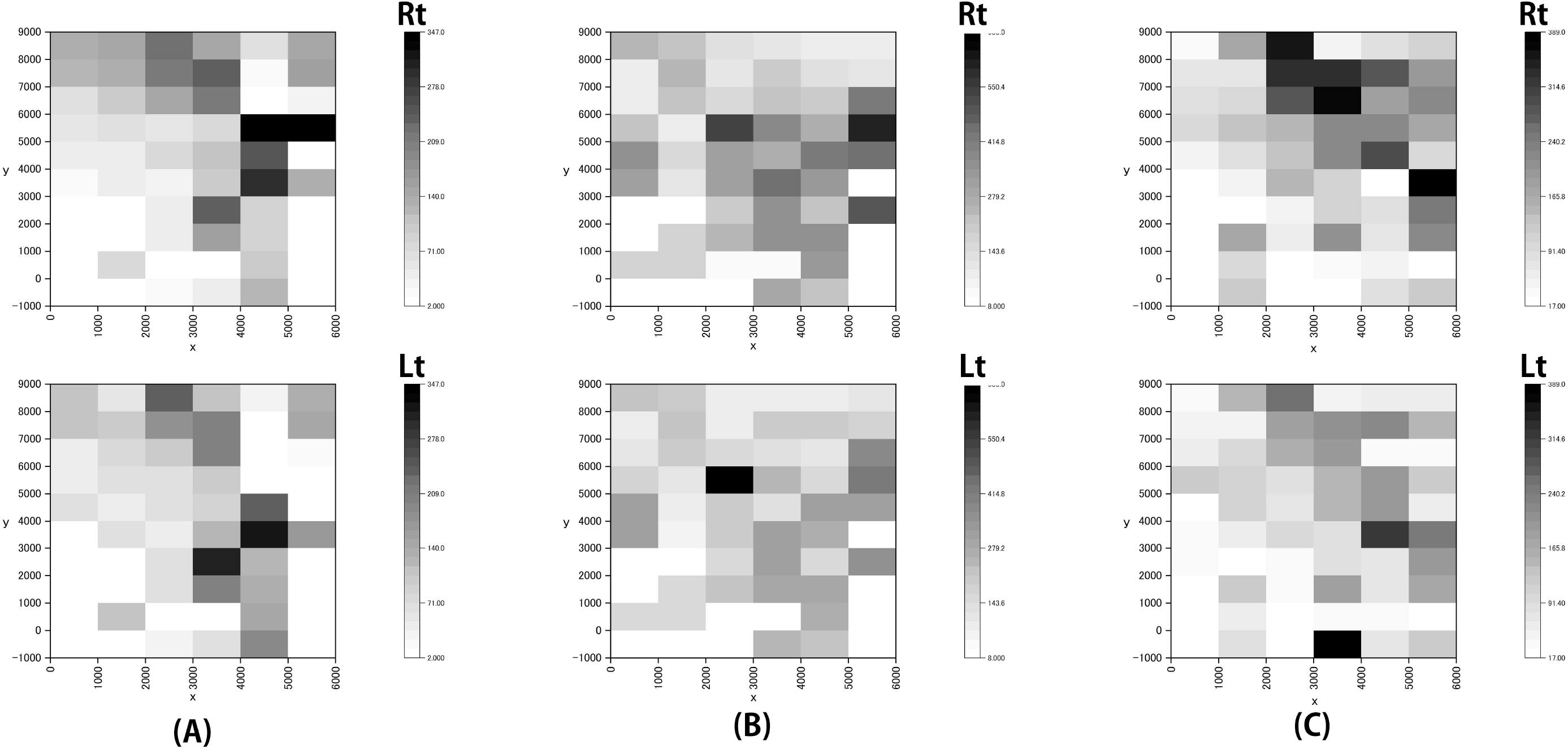
Heat maps. The color of each area indicates the median dust concentration in that area (Rt, right-hand side; Lt, left-hand side).

## Discussion

To assess the accuracy of each experiment, it was found that the error in this measurement system was approximately 1 m. In a previous study, it was reported that the average measurement error of Wi-Fi RTT was 1.4 m in an outdoor experiment in which the measurement points were set in a grid pattern at equal intervals.^11^ Another study reported the average measurement error of Wi-Fi RTT to be 0.10 m in an outdoor experiment and 0.23 m in an indoor experiment in which the measurement points were set along a straight line with equal intervals.^12^ Although the accuracy in this study was worse than this, it is considered to be good enough for use in the workplace. The accuracy of GNSS positioning using a smartphone has been reported to be approximately 8–10 m,^13^ and Wi-Fi RTT is expected to be more accurate than the GNSS if properly configured.

We were able to obtain the same path as the walker indoors using the Wi-Fi RTT. Although there was a certain amount of error in the accuracy verification, we were able to measure continually the path of movement, and there was no interruption in the path. The experiments were conducted on three different walking routes, and no remarkable differences were observed in the positioning results of Wi-Fi RTT. It was assumed that the human body influenced the Wi-Fi RTT positioning, as it has been reported previously^10^. To overcome this issue, the smartphone was held at a high position. In addition, in this study, we coated the glass and metal parts in advance, which were found to degrade the accuracy of measurements in the preliminary investigation. In this regard, it has been assumed that the reflection and refraction of radio waves caused by glass and metal reduces the accuracy. Hence, it is necessary to conduct tests in advance and resolve this issue before using Wi-Fi RTT for measurements in actual workplaces. There have been attempts to create fingerprints using Wi-Fi RTT and utilize machine learning.^14^ Accordingly, the accuracy of this method is expected to improve as the technology develops. However, most of the validation studies on the accuracy of Wi-Fi RTT have been conducted in empty plazas or ordinary offices, and it is desirable to conduct these verification studies in industrial settings, such as factories. Although we did not verify the accuracy of the measurement while walking this time, it is presumed that there is a certain degree of accuracy even when walking given that the walking path was measured without interruption (Figure 5). Considering the fact that the mesh of the heat map is 1 m, the measurement was conducted by walking at an extremely low speed. In the actual work site, it is assumed that people will move by walking more quickly. Given that most of the work itself is done in stationary conditions, and the accuracy in such cases has been verified, it is not expected to be a major problem. To expand the usage of indoor positioning, an additional study on accuracy during motion is required.

One of the advantages of Wi-Fi RTT is that it can be used in local networks, while the GNSS requires orbital information from satellites to identify the location. Without the Internet, it takes more than 30 s to obtain orbital information from GNSS satellites, and the accuracy is initially low.^15^ Given that Wi-Fi RTT requires only an AP and a smartphone for positioning, it does not require this external connection or orbit information acquisition. Instead, Wi-Fi RTT requires the coordinate information of the AP, which is not a concern as the coordinate information of the AP is already known by the company. This can offer a great advantage in the field of occupational health given that external connections are often difficult to access in companies. In this study, we used Wi-Fi RTT in combination with dust measurement, but the GNSS has already been used in various applications, such as geo-fencing, for disaster prevention.^16^ If highly accurate positioning becomes achievable indoors, various applications can be expected to be possible.

Unlike the existing methods, Wi-Fi RTT calculates the distance based on the arrival time of the radio wave rather than on the strength of the radio wave. Theoretically, the radio wave strength attenuates in proportion to the square of the distance, but in reality, this is not always the case owing to the effects of various noise sources. The actual formula for estimating distance using radio wave strength is as follows^17^:

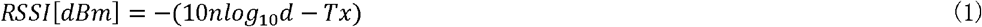

In this formula, RSSI is the strength of the received signal, d is the distance between the transmitter and receiver, n is a constant that represents the attenuation, and Tx is the output of the transmitter. Moreover, n varies depending on the environment; therefore, it is difficult to uniquely determine it in advance. In contrast, Wi-Fi RTT uses the arrival time, and is thus considered to be less susceptible to noise. However, both methods are affected by the reflection, refraction, and diffraction of radio waves; hence, it is necessary to pay attention to this point. In this study, the smartphone was placed on a helmet to reduce these effects. However, considering safety protocols, it may not be realistic to use this method at the workplace. A more stable fixation method should be considered along with technological advances that make the machine smaller and lighter. In this study, we set the height of all the APs and smartphones to be almost the same by ignoring the Z-axis. However, if each AP is set at a higher position such that the radio wave can reach the smartphone directly without being blocked, more stable positioning may be possible; this possibility requires further verification.

Although there is no specific limit to the number of times Wi-Fi RTT can be used, the Android OS specification sets a limit to the number of times the Wi-Fi APs can be scanned.^18^ In addition, there is a limit to the number of times the Wi-Fi RTT can be used in the background and for other location information; however, no such limit exists for its use in the foreground.^19^ It was reported that Wi-Fi RTT consumes excessive battery and is associated with certain privacy issues. When the restriction is enabled, scanning for Wi-Fi APs is suppressed. This may prevent adequate positioning. We eliminated this restriction by using the developer mode in this study, which enabled us to unlock the restriction. While it is possible to consider unlocking the restriction in the same way for company-owned smartphones, this method is not recommended for individually owned smartphones because unlocking is a function for developers. Restrictions may change over time and should be monitored carefully.

In this study, the dust measurement outcomes of the right upper arm were generally higher than those of the left upper arm. The right side of the walking route was closer to the humidifier than the left side, and the results were considered to be consistent with this. Looking over the heat maps of the three experiments, it can be confirmed that the dust concentration near the humidifier with a 5% saline solution (6600, 3600) was higher than in other areas. This can be considered a reflection of the dust emission situation in the experimental environment. In addition, it can be confirmed that the dust concentration near “x = 2000 − 4000, y = 7000 − 9000” is high. In this experiment, the humidifier, which is a dust source, was installed at (0, 6400) and (6600, 3600), and resulted in a gap between the dust source and areas of high concentration in the heat map. One of the reasons for this discrepancy may be the influence of errors in the position measurement. Given that the location information acquisition method used in this experiment had certain limitations, it is possible that the location information was recorded at the point at which the measurer did not actually exist, and the location on the heat map associated with a relatively high-dust concentration was shifted. However, it is possible to interpret the results by assuming that the influence of the error in positional measurement was not significant, i.e., the heat map accurately reflected the dust concentration in the environment to some extent. There could be two possible reasons for this. First, the dust may have been carried away by the movement of the measurer to the location in question. This point was visually confirmed during the experiment (including the preliminary experiment), although it has not been precisely verified or evaluated. Second, there is a possibility of the dust being shifted to the location in question by the operation of the ventilation fan in the simulated factory. By creating a heat map of the dust levels, it was possible to visually understand the dust emissions in the environment. It is possible that the high-dust concentrations formed at a distance from the dust-emission source were not fully identifiable by the usual measurement of the working environment. In this study, we used a 1 m mesh, which was more detailed than the 6 m mesh for measurements in the working environment. Although it was not possible to measure all the points simultaneously, it was still possible to obtain data at many points.

The ability to visually and easily confirm the distribution of dust by combining location information and dust meter results is considered extremely useful in implementing industrial health activities. There have been reports that aimed to gain an understanding on the distribution of the air dose rates after the accident at the Fukushima nuclear power plant to map the concentration/amount of hazardous substances in combination with location information.^20, 21^ However, these reports have used the GNSS in outdoor environments. In the past, indoor positioning was difficult. Therefore, to create these heat maps, it was necessary to determine the position from video camera images, which was very complicated. In addition, personal exposure measurement sometimes involves synchronization of continuous recordings using a real-time monitor with a data logger and video images acquired during work to visually capture the changes in exposure that accompany work changes.^22^ In the past, work scenes were filmed and analyzed, but recent studies have been conducted wherein a small camera and a small dust meter were attached to a helmet, and the camera and dust meter were synchronized to assess the dust emission status.^23^ Although this approach has been in use for a while, it has not been extensively used in actual occupational health settings. One of the reasons for this is attributed to the fact that as the measurement time increases, more time is required to check the video images during the analysis of continuous recordings of the exposure status and the synchronization of the video images. Therefore, it would be more practical to focus on situations associated with high exposures rather than to check all the situations during the entire measurement period. Our method had a great advantage in that we could overview the entire workplace in a relatively short time. However, with the use of our study’s method, if a worker stays in the same place for a long time, the heat map will show only one point; hence, it is necessary to limit the analysis duration or adjust the size of the heat map. In case it is not possible to uniquely link the work location with the work content, a combination of video images can be considered.

This study had a few limitations that must be considered. First, measurement in the study was conducted using only one piece of equipment. It was not clear whether measurements obtained from this equipment can be generalized to those obtained from other equipment. However, at present, few devices officially support Wi-Fi RTT in Japan. It is also unavoidable that the most commonly available devices are made by Google. In particular, there are only four APs listed on the Android developer site as Wi-Fi RTT-compliant APs, and three of them are made by Google.^24^ The fourth one is made by a company other than Google, but it is a foreign product and cannot be used because it does not have a technical standards compliance certificate in Japan. However, the number of compatible smartphones is gradually increasing, and we expect that this number will continue to increase. Second, we did not conduct simultaneous measurements of the measurer’s position using the standard method. Although no large discrepancies between the movement trajectories are presumed, it was not possible to quantitatively evaluate the degree of discrepancy. Given that the accuracy required for use in the workplace is likely to vary from workplace to workplace, additional evaluations are necessary. Third, the simulated factory used in this study had fewer obstructions than a typical factory, and hence, the measurement accuracy may have been overestimated. It is necessary to conduct surveys in more environments, and in certain cases, to combine methods such as fingerprinting for more robust results.

High-precision indoor positioning technology is expected to be used in a wide range of applications, such as the monitoring and evaluation of the working environment and the vital signs of workers, based on its combination with various sensors. Furthermore, this technology is of value in the field of occupational health as it can be used to conduct evaluations in various environments and with various measurement instruments.

## Conclusion

In this study, we visually clarified the distribution of dust concentration in the workplace in the form of a heat map by combining Wi-Fi RTT and WPM. The results in this study indicate that the combination of location information and various sensor devices may be beneficial for occupational health. Additional developments in this forefront are expected in the future.

## Data Availability

The data that support the findings of this study are available from the corresponding author upon reasonable request.

## Acknowledgements

We would like to thank Dr. Takashi Moritake, Dr. Yo Ishigaki, and Mr. Norihisa Kosukegawa for their efforts in the development of the WPM used in this study. We would like to thank Editage (http://www.editage.com) for editing and reviewing this manuscript for English language. This study was funded by the Industrial Disease Clinical Research Grant (180302-01).

## Disclosures

Approval of the research protocol: N/A (All measurements in this study were performed by the authors and no subjects were involved. Therefore, the study was not reviewed by the Ethics Committee because it was not considered to meet the requirements for ethical review.)

Informed Consent: N/A

Registry and the Registration No. of the study/trial: N/A

Animal Studies: N/A

Conflict of Interest: Authors declare no conflict of interest for this article.

## Figure Legends

**Appendix 1.**
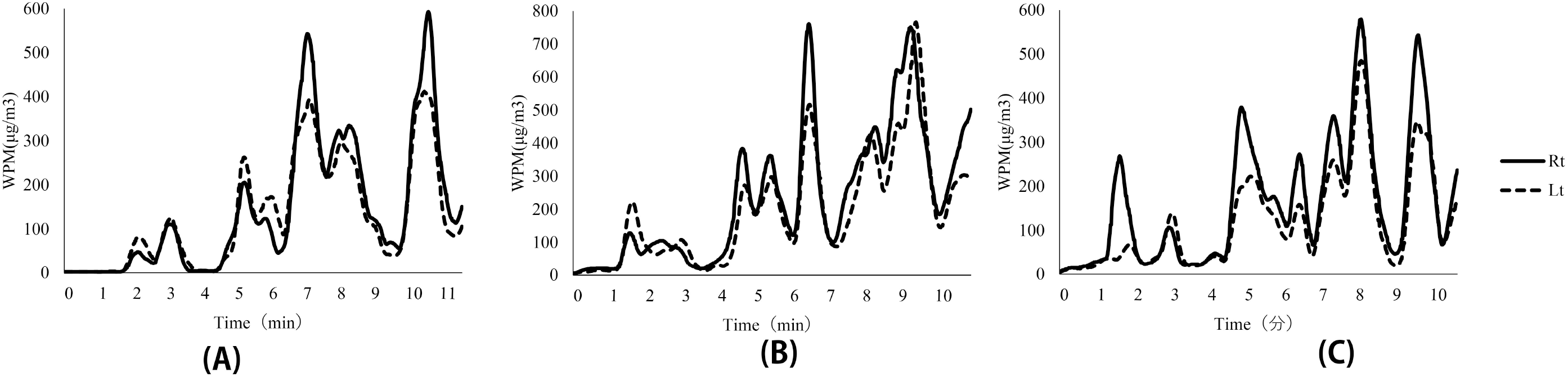
Results of dust measurements (Rt, right-hand side, Lt, left-hand side).

